# Predicting clinical needs derived from the COVID-19 pandemic: the case of Spain

**DOI:** 10.1101/2020.04.03.20051821

**Authors:** Luis Angel Hierro, Antonio José Garzón, Pedro Atienza, José Luis Márquez

**Affiliations:** Departamento de Economía e Historia Económica. Universidad de Sevilla; Hospital Virgen del Rocio. Sevilla

**Keywords:** COVID-19, predictions, clinical needs, ICU, mortality

## Abstract

**Background:** The evolution of the pandemic caused by COVID-19, its high reproductive number and the associated clinical needs, is overwhelming national health systems. We propose a method for predicting both ICU requirements and the number of deaths, and which will enable the health authorities of the countries involved to plan the resources needed to face the pandemic as many days in advance as possible.

**Methods:** We use official Spanish data to predict ICU admissions and deaths based on the number of infections. We employ OLS to perform the econometric estimation, and through RMSE, MSE, MAPE, and SMAPE forecast performance measures we select the best lagged predictor of both dependent variables.

**Findings:** For Spain, our prediction shows that the best predictor of ICU admissions is the number of people infected eight days before, and that the best predictor of deaths is the number of people infected five days before. In the first case, we obtain a 98% coefficient of determination, and in the second a 97% coefficient. The estimated delayed elasticities find that a 1% increase in the number of cases today will imply a 0.72% increase in ICU patients eight days later and a 1.09% increase in the number of deaths five days later.

**Interpretation:** The model is not intended to analyse the epidemiology of COVID-19. Our objective is rather to estimate a leading indicator of clinical needs. Having a forecast model available several days in advance can enable governments to more effectively face the gap between needs and resources triggered by the outbreak and thus reduce the deaths caused by COVID-19.

**Research in context:** *Evidence before this study:* Daily news regarding the exponential growth of those affected by COVID-19 shows that healthcare resources are being overwhelmed by clinical needs in many countries. In particular, serious problems are arising in the most affected countries due to the shortage of ICU beds and the large number of deaths that the authorities are unable to deal with. National health authorities do not have adequate prediction mechanisms to facilitate clinical crisis management. We have performed bibliographic searches of the usual terms used to designate COVID-19, together with those of “prediction”, “estimation”, “ICU”, “mortality” and the like, both in Pubmed and in Google Scholar. The predictive literature related to COVID-19 remains very sparse and the few models that do exist are based on exponential adjustments for forecasting the population affected. However, these models lose their predictive accuracy when the growth rate of infections decreases, added to which such models fail to determine the most statistically efficient maximum prediction time.

*Added value of this study:* We apply a previously unused method based on predictions through delayed logarithmic estimates of ICU admissions and deaths based on the number of infections. For Spain, we estimate that the best predictor of ICU admissions is the number of people infected eight days before and that the best predictor of deaths is the number of those infected five days before. The findings herald a step forward that improves the possibility of managing the health crisis.

*Implications of all the available evidence:* We provide a method to estimate a leading indicator of needs, which thus far has been unavailable to health authorities and which should allow them to plan for the resources required. Furthermore, it is a versatile and simple method that is applicable to any country, state, region, city or hospital area as well as to any type of health care need associated with the COVID-19 pandemic and similar future ones.

## Introduction

The global health crisis sparked by the COVID-19 pandemic has alerted governments worldwide. The speed of transmission caused by the high reproductive number of COVID-19^1^ and the clinical needs associated with the clinical management of patients suffering from severe acute respiratory infection (SARI)^2^ has led to a gap between resources and needs which may prove decisive to the survival of many patients.

COVID-19 entails a great need for clinical treatment, particularly extracorporeal membrane oxygenation (EMO) to patients with acute respiratory distress syndrome (ARDS), and which is pushing existing ICU facilities to breaking point as the pandemic expands^3^. As contagion worsens in each country, the best short-term strategy for containing the spread and so preventing health services from collapsing is social distancing, principally through a quarantine imposed by confining the population, as more and more governments are gradually doing, following WHO recommendations^4^. The objective of the WHO and of national authorities is to flatten the epidemic curve^5^ so that health systems retain the capacity to clinically attend to patients in hospital centres and ICUs, while the scientific community strives to develop an effective vaccine or a viable antiviral.

Increasing health resources rapidly is both complex and costly. The first country to be affected, China, promoted the building of macro-hospitals. Spain and Italy, the two countries currently most affected, have tried to drastically reduce time by using existing macro-buildings such as conference and trade fair centres. This strategy reduces the cost and, particularly, the time needed to set up ICU beds, making the volume of resources required depend mainly on the supply of medical equipment and on the availability of healthcare workers (Spain set up a field hospital with hundreds of beds at the IFEMA fair and conference facility in under 24 hours, and has since continued to expand it to an expected 5.000 beds (https://elpais.com/sociedad/2020-03-21/llegan-los-primeros-pacientes-al-hospital-abierto-en-los-pabellones-de-ifema.html).

Another issue that countries are having to face is how to deal with COVID-19 deaths. In countries where the mortality rate is higher, local funeral systems are unable to meet the cremation needs of COVID-19 deaths, posing saturation problems in the mortuaries. Italy has employed the army to distribute the corpses throughout the territory, moving them away from the most affected areas (http://www.rainews.it/dl/rainews/media/Coronavirus-troppi-morti-a-Bergamo-esercito-porta-le-bare-in-altre-citta-9987ef6c-1056-47c4-862c-18e9255a541a.html#foto-1) while the authorities in Madrid have converted an ice rink into a makeshift mortuary (https://elpais.com/espana/madrid/2020-03-23/madrid-utilizara-como-morgue-las-instalaciones-del-palacio-de-hielo.html).

In these circumstances, where time is key to determining how many lives might be saved, it is essential to have predictive models which can give health authorities a few days prior warning so as to expand their resources and adapt them to the needs they will soon be facing. Our work seeks to provide a predictive model that allows us to determine the needs of ICU beds as far in advance as possible, since admission to ICU offers around a 50% chance of survival for critically ill patients^9^. The model also helps to deal with the practical arrangements involved in dealing with so many deaths.

## Methodology

Through its Ministry of Health website (https://www.mscbs.gob.es/), the Spanish government is providing daily data on the number of detected cases of infected people, deaths, and ICU admissions nationwide. Using officially published data, we econometrically estimate the following equations:

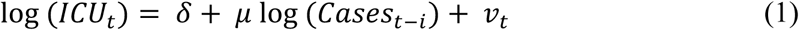

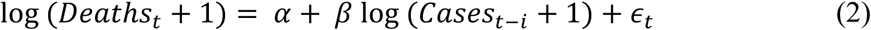

where i=1, 2 …,10 are the number of delays of the explanatory variable, *Deaths*_*t*_ is the total number of deaths up to day t, *Cases*_*t*_ is the number of cases detected up to date t, *ICU*_*t*_ is the number of ICU admissions up to date t, and t=1, 2 …,T representing each day. In equation (2), variables are defined as log *(Deaths*_*t*_ + 1) and log *(Cases*_*t-i*_ + 1) to avoid missing observations from the first few days where the number of deaths was zero. For equation (1), the sample period spans from 08/03/2020 to 24/03/2020, while in equation (2) the period spans from 28/02/2020 to 24/03/2020. We left March 25 and 26 as out-of-sample observations in order to measure the forecast performance of the estimated model. The estimation sample is shorter for equation (1) due to the lack of observations. Estimation is performed through the OLS estimator.

Given that our main goal is to predict ICU beds and the number of deaths using the lagged number of infected cases, after estimating equations using different lags, we select the one with the best forecast performance (which minimises forecasting errors). Using data from 25/03/2020 and 26/03/2020 as out-of-sample period, we calculated RMSE, MSE, MAPE, and SMAPE as indicators of predictive capacity. RMSE is defined as follows:

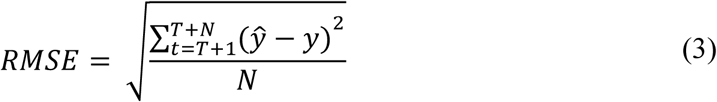

where N=2 is the number of out-of-sample observations, which we use to estimate the forecast performance of our estimate, *ŷ* is the estimated value of the dependent variable, and *y* is the actual value. Secondly, MSE is defined as follows:

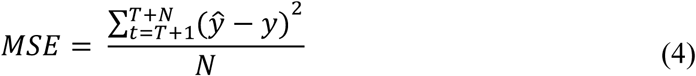

MAPE is calculated using the following formula:

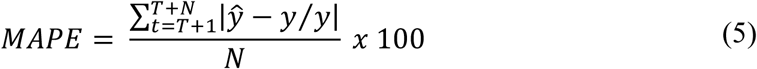

SMAPE follows this formula:

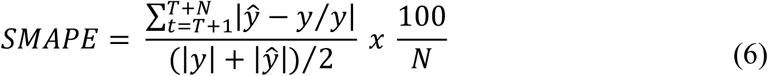

Finally, we select the estimate with the lagged explanatory variable that shows the lowest value in this set of indicators, and we make the corresponding prediction in total values.

## Results

### a. Prediction of patients in ICUs

First, we select the model that has the best forecast performance. Table 1 shows RMSE, MSE, MAPE, and SMAPE values for the estimation of equation (1), using from 1 to 10 delays of the explanatory variable. We select the model including eight delays, since it shows the lowest value in the different indicators, which means it outperforms the other estimates in forecasting accuracy.

**Table 1.**
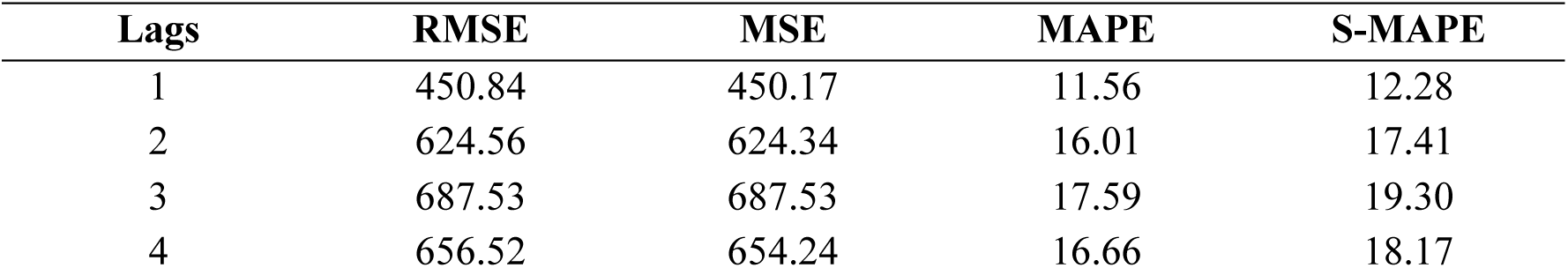
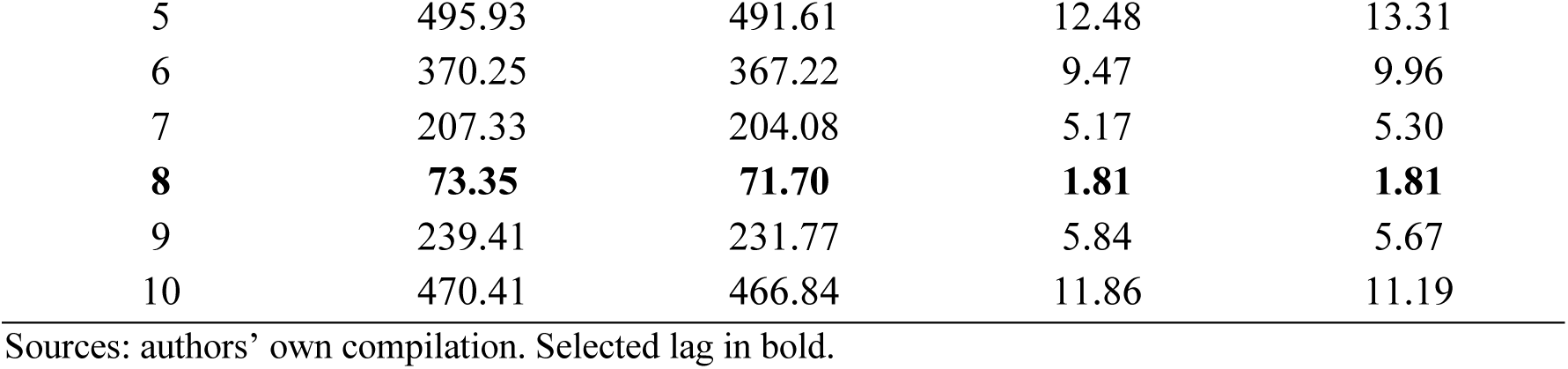
Out-of-sample forecast performance for ICU beds (equation 1).

| Lags | RMSE | MSE | MAPE | S-MAPE |
| --- | --- | --- | --- | --- |
| 1 | 450.84 | 450.17 | 11.56 | 12.28 |
| 2 | 624.56 | 624.34 | 16.01 | 17.41 |
| 3 | 687.53 | 687.53 | 17.59 | 19.30 |
| 4 | 656.52 | 654.24 | 16.66 | 18.17 |
| 5 | 495.93 | 491.61 | 12.48 | 13.31 |
| 6 | 370.25 | 367.22 | 9.47 | 9.96 |
| 7 | 207.33 | 204.08 | 5.17 | 5.30 |
| **8** | **73.35** | **71.70** | **1.81** | **1.81** |
| 9 | 239.41 | 231.77 | 5.84 | 5.67 |
| 10 | 470.41 | 466.84 | 11.86 | 11.19 |

Sources: authors' own compilation. Selected lag in bold.

The equation of the model that evidences the best forecast performance is the following:

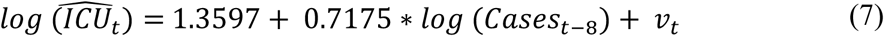

The coefficient *μ* = 0.7175 is what in economics is called elasticity; in this case delayed. We obtain it by deriving the function, and it represents the relationship between the variation of the dependent and independent variables:

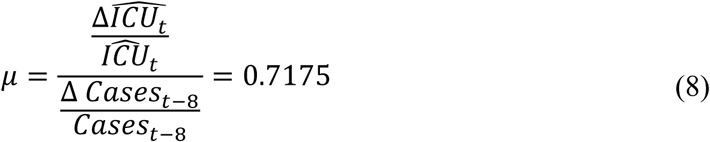

This delayed elasticity implies that a 1% increase in the number of infected cases predicts a 0.72% increase in the number of ICU patients eight days later. The estimate presents an R-square of 0.98.

Our model allows us to predict the number of ICU beds to be used, taking the number of people infected eight days earlier as a reference. In this way, we can anticipate the needs in terms of the number of beds for the next few days.

Figure 1 displays the estimates of equation 1 compared to the actual values, expressed in logs, as well as the evolution of the residuals over time. As can be seen, the residuals decrease throughout the period; that is, the estimate of the number of ICU beds is closer to the actual values.

**Figure 1.**
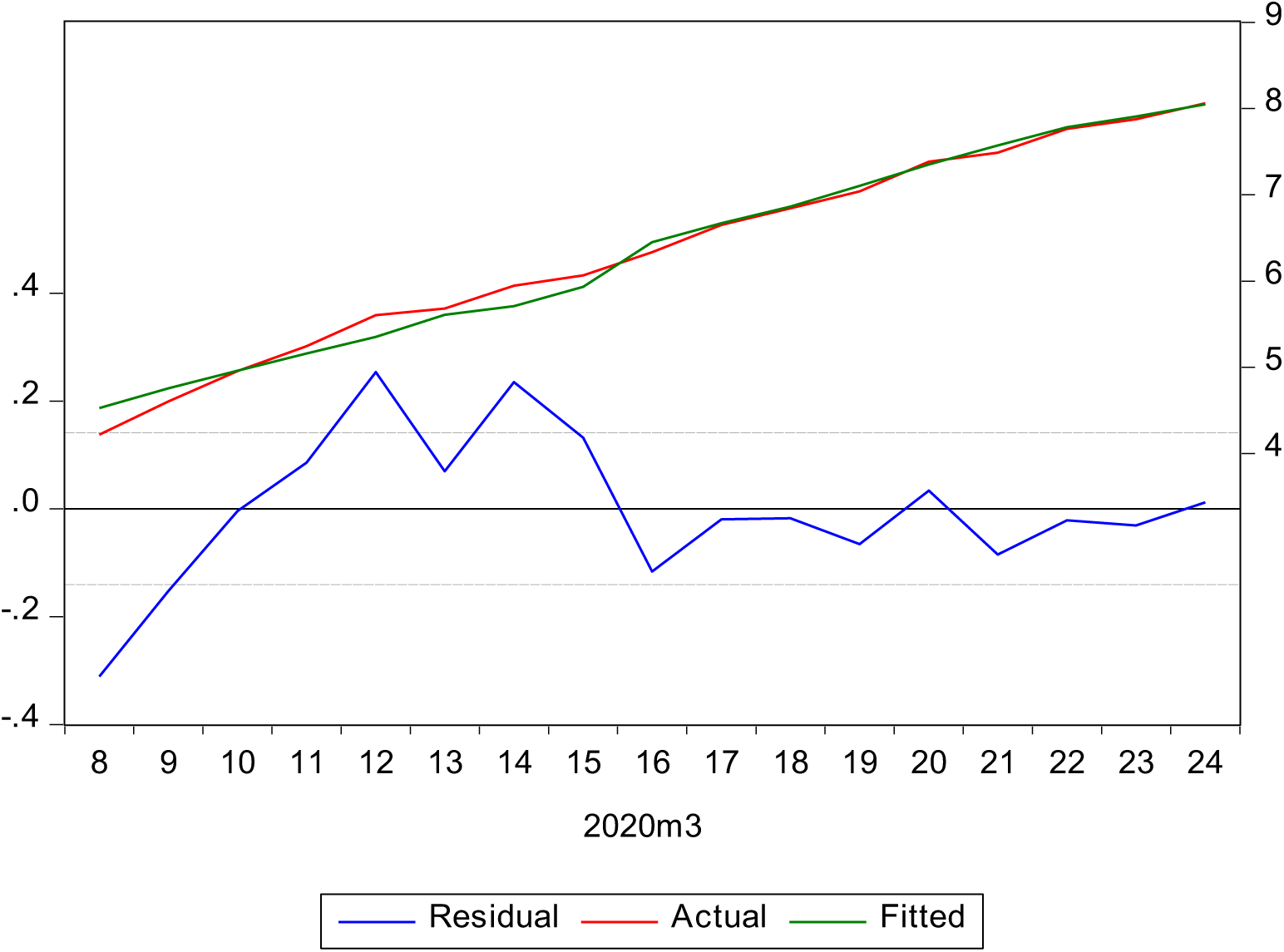
Model estimate values, actual values, and residuals (equation 1) Source: authors’ own compilation

Table 2 shows the actual and estimated total number of ICU admissions for the period covered by the estimation, as well as the estimation error of the model. In addition, we include out-of-sample observations, which allows us to analyse the forecast performance of the model. To obtain the actual total number of ICU admissions, we perform the following transformation:

**Table 2.** Actual vs estimated ICU admissions and estimate error

| Date | Cases | ICU | ICU (Est) | Error | Error (%) |
| --- | --- | --- | --- | --- | --- |
| 08/03/2020 | 1204 | 68 | 93 | 24.79 | 36.45 |
| 09/03/2020 | 1639 | 100 | 117 | 16.51 | 16.51 |
| 10/03/2020 | 2140 | 142 | 143 | 0.55 | 0.39 |
| 11/03/2020 | 3004 | 190 | 174 | -15.60 | -8.21 |
| 12/03/2020 | 4231 | 272 | 211 | -60.90 | -22.39 |
| 13/03/2020 | 5753 | 293 | 273 | -19.73 | -6.73 |
| 14/03/2020 | 7753 | 382 | 302 | -79.96 | -20.93 |
| 15/03/2020 | 9191 | 432 | 379 | -53.46 | -12.37 |
| 16/03/2020 | 11178 | 563 | 632 | 69.28 | 12.31 |
| 17/03/2020 | 13716 | 774 | 789 | 14.90 | 1.93 |
| 18/03/2020 | 17147 | 939 | 955 | 16.30 | 1.74 |
| 19/03/2020 | 19980 | 1141 | 1218 | 77.48 | 6.79 |
| 20/03/2020 | 24926 | 1612 | 1558 | -54.09 | -3.36 |
| 21/03/2020 | 28572 | 1785 | 1942 | 157.21 | 8.81 |
| 22/03/2020 | 33089 | 2355 | 2406 | 50.85 | 2.16 |
| 23/03/2020 | 39673 | 2636 | 2718 | 82.25 | 3.12 |
| 24/03/2020 | 47610 | 3166 | 3128 | -37.92 | -1.20 |
| **25/03/2020** | **56188** | **3679** | **3623** | **-56.24** | **-1.53** |
| **26/03/2020** | **64059** | **4165** | **4252** | **87.17** | **2.09** |
| **27/03/2020** |  |  | **4745** |  |  |
| **28/03/2020** |  |  | **5561** |  |  |
| **29/03/2020** |  |  | **6134** |  |  |
| **30/03/2020** |  |  | **6815** |  |  |
| **31/03/2020** |  |  | **7763** |  |  |

Source: authors' own compilation and Ministry of Health. ICU(est) values are rounded to integer values. Out-of-sample dates in bold.

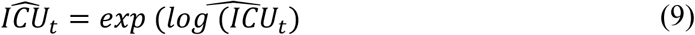

That is, we convert the values in logs to total numbers. As the results show, the estimation error, measured as a percentage of total ICU admissions, decreases over the period, although in absolute terms it is greater. Figure 2 graphically shows these same results, including the ICU number estimated until 31/03/2020. As can be seen, the model allows us to very accurately predict the number of ICU patients.

**Figure 2.**
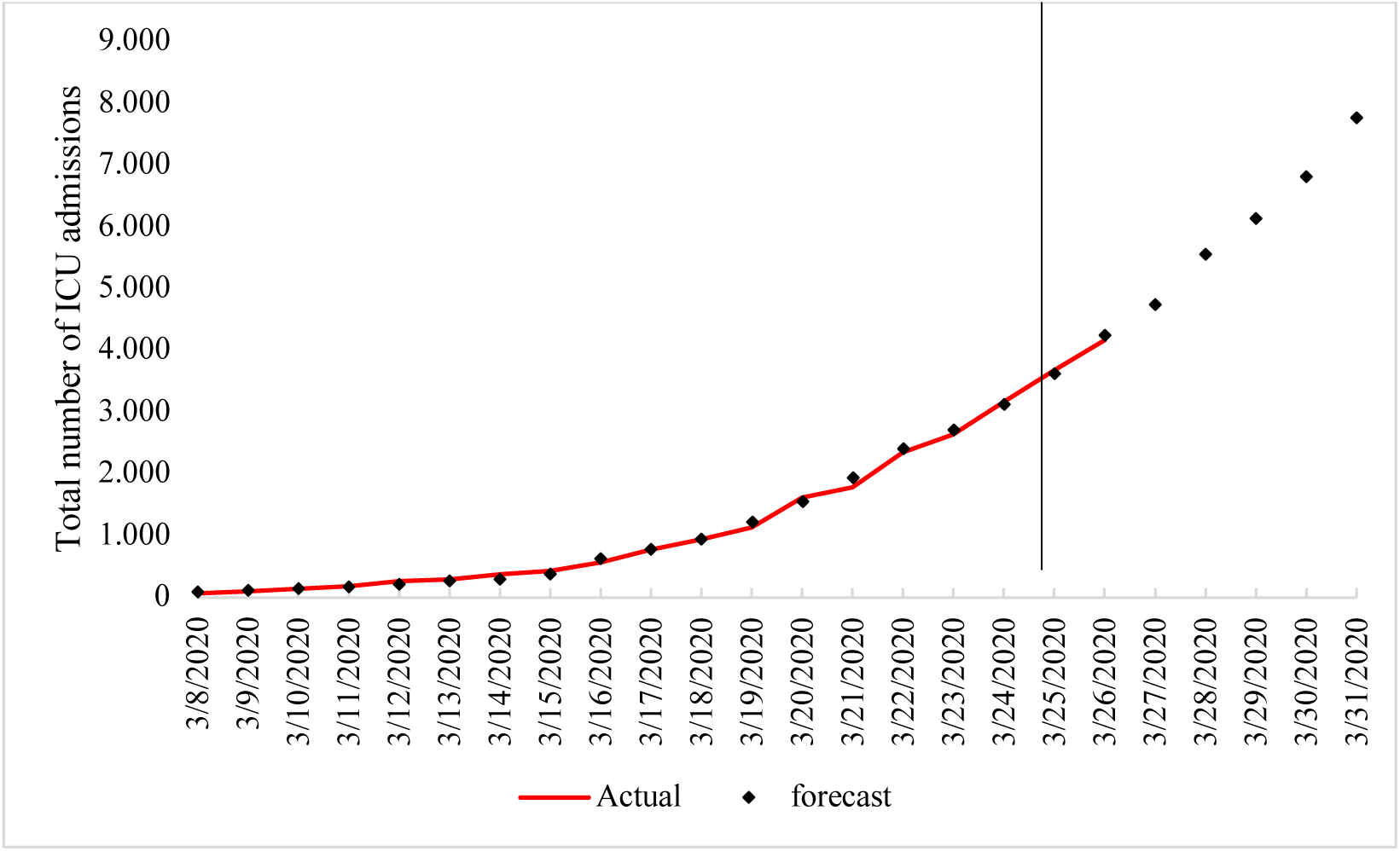
Actual vs estimated total ICU admissions. Source: authors’ own compilation and Ministry of Health

### b. Prediction of deaths

We proceed in the same way in the case of deaths caused by COVID-19. Table 3 shows the different forecast performance indicators calculated for the different models in equation (2) using up to 10 delays. According to the estimates, we select the model that includes the explanatory variable with five delays, since it produces the lowest values for the four different measures of forecast performance (RMSE, MSE, MAPE, and SMAPE).

**Table 3.** Out-of-sample forecast performance for COVID-19 deaths (equation 2).

| Lags | RMSE | MSE | MAPE | S-MAPE |
| --- | --- | --- | --- | --- |
| 1 | 230.1 | 222.15 | 5.12 | 5.27 |
| 2 | 205.5 | 182.51 | 4.29 | 4.42 |
| 3 | 268.34 | 260.13 | 5.99 | 6.19 |
| 4 | 207.06 | 205.05 | 4.56 | 4.69 |
| **5** | **84.31** | **62.17** | **1.29** | **1.31** |
| 6 | 174.02 | 145.77 | 3.1 | 3.04 |
| 7 | 453.61 | 453.58 | 10.2 | 9.7 |
| 8 | 1108.6 | 1079.67 | 23.82 | 21.25 |
| 9 | 1808.13 | 1787.04 | 39.71 | 33.11 |
| 10 | 2833.87 | 2811.38 | 62.62 | 47.68 |

Source: authors' compilation. Selected lag in bold.

We select the five-day delayed model, since it shows the best forecast performance according to the indicators.

The equation of the model that presents the best forecast performance is:

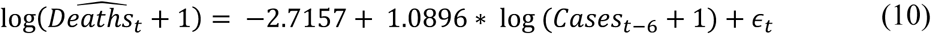

The delayed elasticity function is as follows:

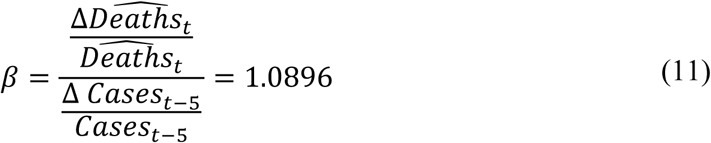

showing that a 1% increase in the number of infected cases detected predicts a 1.09% increase in the number of deaths six days later. The estimate presents a high goodness of fit, with an R-square of 0.97.

Figure 3 displays the estimate of the model with respect to the actual values, both in logs. As was the case with ICU numbers, we find that residuals decrease throughout the estimation period, so that the estimate values at the end of the sample are closer to the actual values.

**Figure 3.**
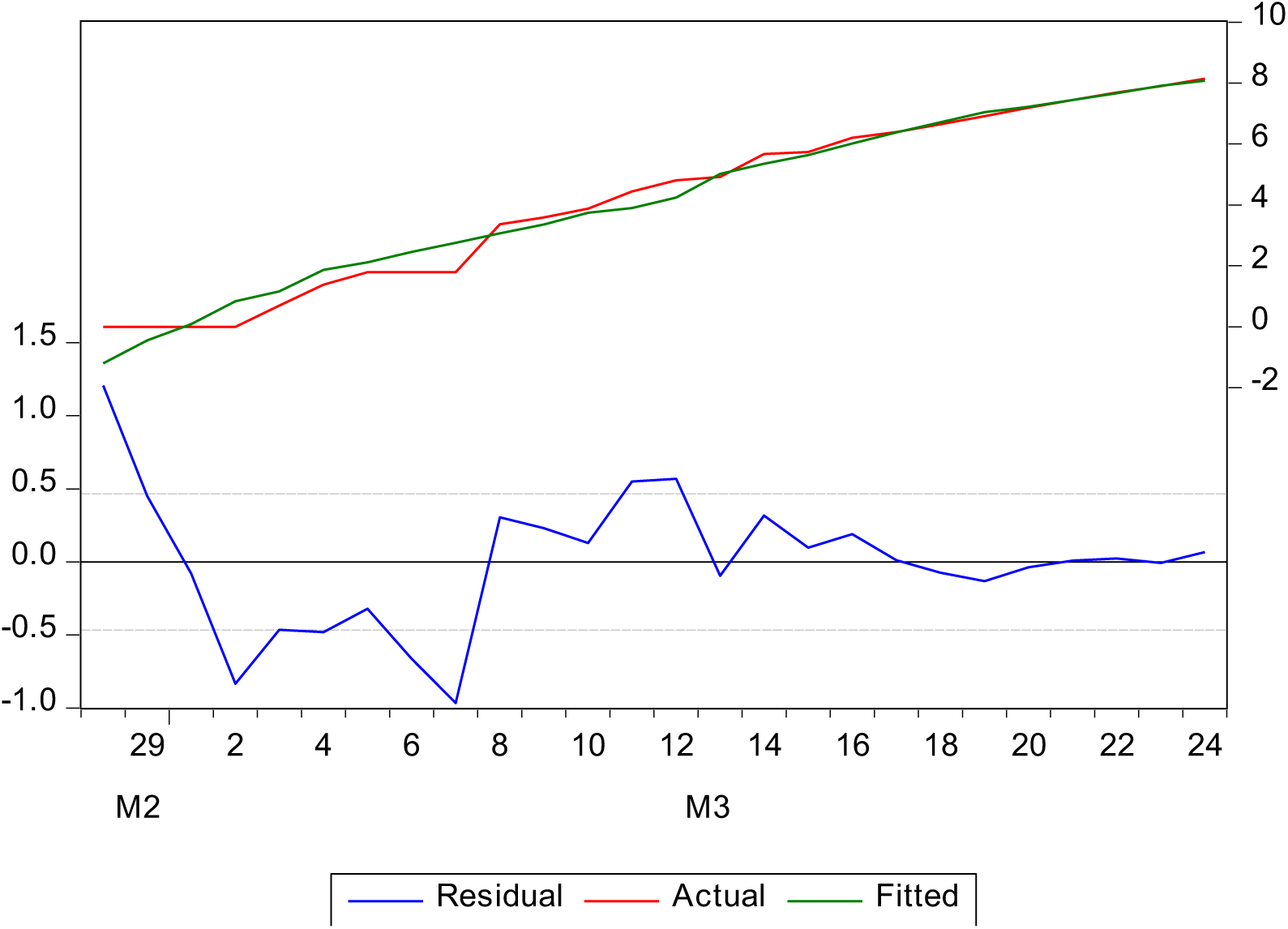
Model estimate values, actual values, and residuals (equation 2) Source: authors’ own compilation

Table 4 shows the number of actual and estimated deaths, as well as the errors for each time period. To obtain the total number of deaths, we carry out the following transformation:

**Table 4.** Actual vs estimated deaths and estimate error.

| Date | Cases | Deaths | Deaths (Est) | Error | Error % |
| --- | --- | --- | --- | --- | --- |
| 08/03/2020 | 1204 | 28 | 20 | -7.61 | 27.19 |
| 09/03/2020 | 1639 | 35 | 28 | -7.45 | 21.29 |
| 10/03/2020 | 2140 | 47 | 41 | -5.81 | 12.35 |
| 11/03/2020 | 3004 | 84 | 48 | -35.90 | 42.74 |
| 12/03/2020 | 4231 | 121 | 68 | -52.87 | 43.69 |
| 13/03/2020 | 5753 | 136 | 150 | 13.53 | 9.95 |
| 14/03/2020 | 7753 | 288 | 210 | -78.40 | 27.22 |
| 15/03/2020 | 9191 | 309 | 281 | -28.42 | 9.20 |
| 16/03/2020 | 11178 | 491 | 406 | -84.59 | 17.23 |
| 17/03/2020 | 13716 | 598 | 591 | -7.37 | 1.23 |
| 18/03/2020 | 17147 | 767 | 826 | 58.85 | 7.67 |
| 19/03/2020 | 19980 | 1002 | 1143 | 141.43 | 14.12 |
| 20/03/2020 | 24926 | 1326 | 1377 | 50.51 | 3.81 |
| 21/03/2020 | 28572 | 1720 | 1704 | -16.09 | 0.94 |
| 22/03/2020 | 33089 | 2182 | 2130 | -52.32 | 2.40 |
| 23/03/2020 | 39673 | 2696 | 2716 | 20.43 | 0.76 |
| 24/03/2020 | 47610 | 3434 | 3209 | -224.95 | 6.55 |
| **25/03/2020** | **56188** | **4089** | **4084** | **-5.21** | **0.13** |
| **26/03/2020** | **64059** | **4858** | **4739** | **-119.12** | **2.45** |
| **27/03/2020** |  |  | **5561** |  |  |
| **28/03/2020** |  |  | **6777** |  |  |
| **29/03/2020** |  |  | **8267** |  |  |
| **30/03/2020** |  |  | **9902** |  |  |
| **31/03/2020** |  |  | **11423** |  |  |

Source: authors' own compilation and Ministry of Health. Deaths(est) values are rounded to integer values. Out-of-sample dates in bold.

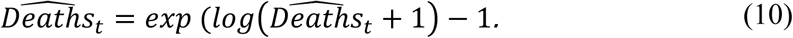

Results show that the prediction error, in percentage terms, decreases over time. As for total values, it increases at the beginning of the period and then subsequently decreases. As regards the predicted out-of-sample values, we observe that the error is very small, with 5 and 119 deaths, which corresponds, in percentage terms, to 0.87% and 2.45%, respectively.

In Figure 4, we see the trend in the actual number of deaths and their estimates in graphical terms. We also include the prediction of deaths obtained by the model up to 31/03/2020. The estimates show a high goodness of fit, presenting an R-square of 0.97, in addition to generating a prediction with a very low error, as shown in Table 3.

**Figure 4.**
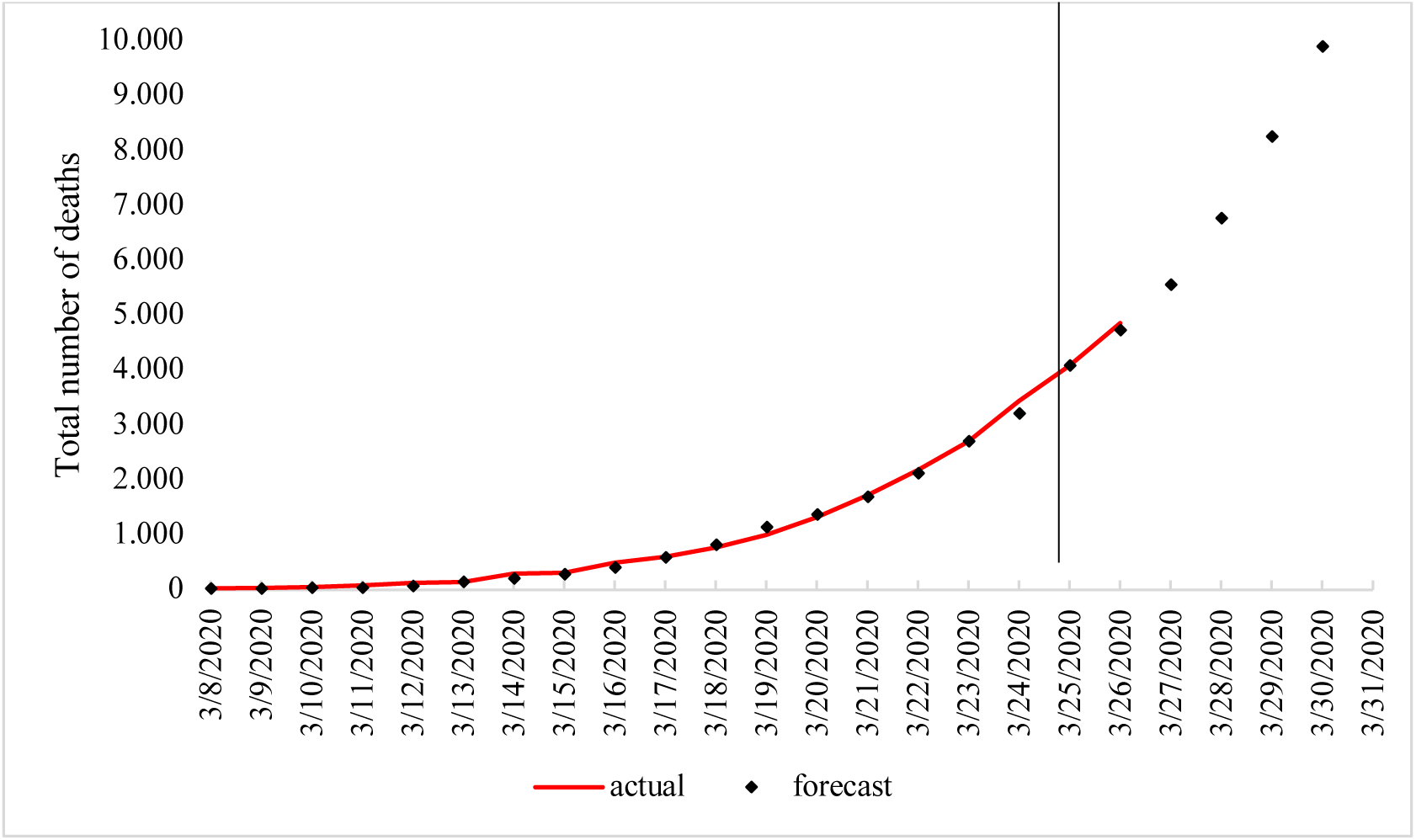
Actual vs estimated total number of deaths. Source: authors’ own compilation and Ministry of Health.

## Discussion

To date, predictive models for COVID-19 have focused on predicting changes from exponential adjustments for the growth in the infected population, without a predefined contagion model^6,7^ or with a predefined contagion model^8^. These types of adjustments are highly dependent on the estimated equation, and lose their predictive accuracy when the growth rate of infections decreases. They also fail to determine the most statistically efficient maximum prediction time^6^. This makes them ineffective for prediction and prevents health authorities from obtaining reliable forecasts.

Our strategy consists of performing simple delayed regressions of the number of patients affected in ICUs, and deaths and by selecting the equation corresponding to the delay that provides the most statistically efficient prediction. By foregoing any epidemiological, or indeed any other type of objective, and by avoiding predetermining the function which is to be estimated, we are able to focus solely and exclusively on finding the best predictor.

The result of this simple strategy is a model with a very high predictive accuracy, offering an eight-day advance for ICU admissions and a five-day advance for deaths. This advanced forecast is what can allow authorities to improve resource planning to face the clinical needs-health resources gap caused by the epidemic.

Clearly, the findings are valid for Spain but not for other countries. However, the method is replicable to any country, state, or region. In addition, it is applicable to any type of health needs, provided we have data on their consumption. It can be applied to anticipate the needs of health staff, hospital beds, medicine, equipment …, and can also be applied to anticipate other clinical situations apart from deaths.

The method does, nevertheless, evidence certain shortcomings that must be taken into account and that basically affect the available data. The method only uses one independent variable, namely those officially declared as being infected, and which thus relies on the possibility of actually applying detection tests. It also depends on health authorities being truthful in their information policy. In Spain, few detection tests were carried out in the early stages of the outbreak, such that patients displaying mild symptoms were not tested. Large-scale testing of mild cases began around 20/03/2020 (https://www.lavozdegalicia.es/noticia/pontevedra/2020/03/20/test-rapidos-salir-coche-inician-hoy-cita-previa/0003_202003P20C1992.htm), although not throughout the whole country. Obviously, if the number of tests for possible COVID-19 infected cases were to be increased, the model would tend to overestimate ICUs and deaths and would need to be recalibrated. In other words, any significant change in the number of tests would affect our independent variable, the number of those declared as being infected, and would have consequences for the predictive performance of both independent variables.

Moreover, a second issue arises in one of the dependent variables. The number of patients in the UCIs represents the actual number of patients there, but not the number of patients who are in need of intensive care. If ICUs are swamped, medical staff will be forced to ethically screen critical patients^10^, such that data will cease to be representative of clinical needs and will become representative of the limited resources available. If we consider that this need to screen will increase the more the pandemic spreads and the greater the number of patients in need of ICU care, the model may be underestimating ICU needs as infection expands. Moreover, any improvement in clinical pharmacological treatment using antivirals in the pre-ICU admission stage will entail an overestimation of needs.

A third problem concerns the communication of data. Given the high reproductive number of COVID-19, contagion is growing exponentially, and having accurate daily data available is essential. When 800 patients die every day, at a rate of 33 deaths per hour, reporting the number of deaths inaccurately can lead to fundamental errors in the estimates. The health authorities must establish a specific time to report the numbers and must keep this stable, added to which health centres must prepare the counts accurately and ensure that they reflect the established time periods. In Spain, it was decided to close the daily count at 9.00 pm, and this is a time which must remain fixed until the pandemic is over. Obviously, public information must be truthful and any cases of UCIs being overwhelmed must be reported.

The effect of data issues can be corrected in two ways: by increasing the variables whose numbers are gathered and published (for example, by publishing daily figures of the tests carried out and of their results or through the numbers of patients not admitted to UCIs as a result of previous ethical screening), and by recalibrating estimates if they lose their predictive accuracy. The first alternative would enable us to replace the variables or to correct them prior to performing estimations. The second alternative would allow us to refine the predictions recursively when the model loses its predictability. Whatever the case, this latter alternative can be performed daily given the ease of the method and the small amount of additional information required.

On a final note, it should be remembered that our work does not pursue any purpose regarding epidemiological analysis, but merely seeks to offer a predictive method which can enhance the planning capacity of health authorities, who are facing the greatest global health challenge the world has seen in the last 100 years.

## Data Availability

all data are available online

